# Evaluation of AI-assisted summarisation of tertiary clinical genomics reports: results of the QNOMX-VHIR-CPSP-001 Phase 1 study

**DOI:** 10.64898/2026.09.10.26362656

**Authors:** James Creeden, Marcus Olivecrona, Nuria Benavent, Raquel Hladun Alvaro, Lorena Valero-Arrese, Asbleidy Carolina Torres, Aroa Soriano

**Affiliations:** Qnomx AG, Basel, Switzerland; Childhood Cancer and Blood Disorders Group, Vall d’Hebron Institut de Recerca (VHIR), Universitat Autònoma de Barcelona, Barcelona, Spain; Paediatric Oncology and Haematology Department, Vall d’Hebron University Hospital, Barcelona, Spain; Clinical and Translational Cancer Research Group, Health Research Institute Hospital la Fe (IIS La Fe), Valencia, Spain

**Keywords:** clinical performance study, ISO 20916, AI-assisted summarisation, clinical genomics, tertiary genomics reports, content fidelity, inter-rater reliability, non-inferiority, precision oncology, Bayesian hierarchical model

## Abstract

**Background:** Comprehensive genomic reports in oncology contain complex molecular information that must be translated into concise summaries for treating clinicians. Manual summarisation is time-intensive, and robust evidence supporting AI-assisted summarisation systems remains limited. We evaluated whether AI-generated summaries were non-inferior to manually produced summaries in faithfully representing information from tertiary paediatric cancer genomics reports.

**Methods:** QNOMX-VHIR-CPSP-001 Phase 1 was a single-site, non-interventional clinical performance study conducted at Vall d’Hebron Institut de Recerca under ISO 20916:2019. De-identified tertiary paediatric cancer genomics reports were summarised using both the AI-assisted system under evaluation and the standard manual workflow. Three qualified raters assessed both summary types against the source reports using the six-domain, five-point Quality Summary Index (QSI) in a blinded, counterbalanced crossover design with a minimum fourteen-day washout. Co-primary endpoints were composite content and presentation scores. Non-inferiority was assessed using a pre-specified Bayesian hierarchical model with a margin of 0.5 points and confirmed using a frequentist linear mixed model.

**Results:** Thirty-seven of 38 planned cases contributed 74 paired evaluations. Mean content scores were similar between AI-generated and manual summaries (4.01 vs 3.97), whereas AI-generated summaries achieved higher presentation scores (4.39 vs 4.12). Non-inferiority was demonstrated for both co-primary endpoints: content +0.05 (95% CrI −0.18 to +0.28), presentation +0.28 (+0.10 to +0.47). Frequentist analyses were concordant. Accuracy was the only QSI dimension for which Bayesian non-inferiority was not concluded. Inter-rater reliability was low across all QSI domains.

**Conclusions:** AI-generated summaries were non-inferior to manually produced summaries for both content and presentation endpoints in tertiary paediatric oncology genomics reports. Presentation scores were higher for AI-generated summaries, whereas content scores were comparable between approaches. Low inter-rater reliability limits confidence in the dimension-level estimates and is the main constraint on future validation studies. Clinical validity and workflow effect were not assessed.

## Introduction

Precision oncology depends on tertiary genomics reports: integrated molecular documents consolidating sequencing results, copy-number findings, fusion calls, and the variant classifications and therapeutic associations attached to them. These reports are dense, often issued in English, and follow reporting standards written for specialist molecular pathology and tumour board use [1,2], but the community oncologist who must act on them at the point of care needs a concise, accurate summary. Today that summary is produced by hand and often translated into local language, a task that is time-consuming and whose outputs vary in completeness and clarity.

Systems that assist this summarisation are now technically feasible, but the evaluation methodology has lagged behind the tools. Human-evaluation studies of large language models in healthcare are frequently criticised for absent sample-size justification, under-specified rater numbers and training, and metrics chosen without a psychometric basis [3,4,5]. When a tertiary report is condensed, several types of error can occur. Clinically relevant findings may be omitted (completeness), unsupported statements may be introduced (accuracy, or “hallucination”), and information may be presented in ways that reduce its usefulness for the intended reader (relevance, organisation, language, and succinctness). These dimensions reflect how faithfully a summary represents its source report, rather than properties of the summary in isolation. We therefore evaluate content fidelity as a co-primary endpoint, defined as the extent to which a summary represents its source without material omission or distortion, following the faithfulness construct developed for long-form clinical summarisation [6,7]. Summaries were assessed against a fixed reference by qualified raters under blinded conditions.

The methods and endpoints were designed to isolate the summarisation task itself. Variant classifications are already established in the source report; evaluating classification accuracy would therefore reflect the performance of the upstream laboratory pipeline, where AI systems for variant-evidence curation are in development and early evaluation [8,9], rather than the summarisation process. Likewise, clinical decision-making outcomes were not appropriate endpoints for this phase of evaluation, since the study was non-interventional and did not assess real-world clinical use. For these reasons, the study focused on content fidelity and presentation quality as measures directly attributable to the summarisation system.

We report the results of QNOMX-VHIR-CPSP-001 Phase 1, which evaluated the content fidelity and presentation quality of AI-assisted summaries of tertiary paediatric oncology genomics reports compared with the manual standard of care. The study followed a pre-specified design and analysis plan published separately as a companion methods preprint [10].

## Methods

### Design and oversight

QNOMX-VHIR-CPSP-001 is a non-interventional clinical performance study documented as a Clinical Performance Study Protocol under ISO 20916:2019 (European adoption EN ISO 20916:2024) [11]; the standard governs study conduct and does not assert a regulatory classification of the system under evaluation. Phase 1 was a retrospective evaluation in which de-identified archived tertiary paediatric cancer genomics reports were summarised by the AI-assisted system under evaluation and, in parallel, by the manual standard workflow, and both summary types were scored against the source report. Outputs of the system under evaluation were not used for patient management; the manual standard-of-care summary remained the only clinical artefact. Throughout this report, “AI-assisted” denotes summaries produced by the AI-assisted summarisation system under evaluation and “manual” denotes summaries produced by the standard workflow; these correspond to the system and manual arms of the study protocol. The study was conducted at Vall d’Hebron Institut de Recerca (VHIR), Barcelona, with Qnomx AG, Basel, as sponsor, under a favourable CEIM opinion (reference PR(AMI)318-2025, dated 08 August 2025), including a waiver of additional informed consent for the non-interventional use of de-identified archived research data.

Cases were drawn from tertiary genomics reports of paediatric cancer cases archived by the COMIK and SEHOP-PENCIL projects at VHIR [12]. All reports from patients with relapsed paediatric solid tumours issued on the same report template version were eligible. Forty-six cases were curated, eight forming the familiarisation set and the remainder the analysis set. De-identification was performed by a designated member of the original study team who was not part of the QNOMX-VHIR-CPSP-001 research team, removing all direct identifiers, original study codes, dates, reference numbers, professional names and referring-hospital information, and retaining only clinically relevant genomic findings, biological sex, age, tumour type, and disease status at inclusion. Each case was assigned a new consecutive number with no retained link to the original code, verified before use, and uploaded to the shared study environment for blinded scoring. One case was excluded before data lock because of a source-document error identified independently of the QSI scores.

The design was a blinded, counterbalanced, two-period crossover at the rater–case level. Blinding was implemented at the artefact level through round-specific pseudonyms, so that raters could not infer a summary’s origin or link the two summary types for the same case across periods, and a minimum fourteen-calendar-day washout (enforced by access control in the study environment) separated each rater’s two scoring events for the same case.

### Raters and the QSI instrument

Ratings were performed by three qualified oncologists, each with at least four years of experience in cancer genomics; one rater scored every analysis case and two further raters each scored a complementary, non-overlapping subset, so that each case received two independent ratings. All raters completed a documented in-person training workshop followed by a familiarisation round in which they scored a single set of eight cases, several modified with deliberately introduced errors, using the study’s electronic scoring tool. The familiarisation set was not split by arm and was excluded from all inference.

The Quality Summary Index assesses a genomics summary against its source report across six dimensions, each scored 1 (unacceptable) to 5 (excellent): Accuracy, Completeness, Relevance, Organisation, Language, and Succinctness.

Quality is summarised by two composites defined as dimension means on the 1–5 scale: a content composite (accuracy, completeness, relevance) and a presentation composite (organisation, language, succinctness); an overall composite is the mean of all six dimensions. The QSI is adapted from the Provider Documentation Summarization Quality Instrument (PDSQI-9) and its Physician Documentation Quality Instrument (PDQI-9) antecedent [13,14]. The instrument and its anchors are reported in full in Table 1 of the methods preprint [10].

**Table 1.** (Results). Dimension- and composite-level QSI scores by arm; mean (SD), n = 74 per arm. SD, standard deviation. Δ values are calculated from unrounded data.

| QSI element | Manual, mean (SD) | AI-assisted, mean (SD) | $\Delta$ (AI-assisted – manual) |
| --- | --- | --- | --- |
| **Content composite** | **3.97 (0.92)** | **4.01 (1.03)** | **+0.04** |
| Accuracy | 3.97 (1.52) | 3.80 (1.71) | −0.18 |
| Completeness | 3.76 (1.29) | 3.80 (1.27) | +0.04 |
| Relevance | 4.19 (1.08) | 4.43 (1.09) | +0.24 |
| **Presentation composite** | **4.12 (0.71)** | **4.39 (0.61)** | **+0.27** |
| Organisation | 4.39 (1.07) | 4.65 (0.88) | +0.26 |
| Language | 4.28 (0.77) | 4.15 (0.93) | −0.14 |
| Succinctness | 3.69 (1.22) | 4.38 (1.02) | +0.69 |
| **Overall composite** | **4.05 (0.70)** | **4.20 (0.66)** | **+0.15** |

The training workshop familiarisation round surfaced points of ambiguity in rubric application that informed the scoring guidance used thereafter.

### Analysis sets and pre-specified analysis

The primary analysis used a complete-case per-protocol dataset comprising all eligible cases with a usable source report and complete QSI assessment for both summary types; a rater-by-report row entered the analysis only where all six dimensions were scored. Because one case was excluded on eligibility grounds identified independently of scoring, a pre-specified intention-to-treat sensitivity analysis including this case was also performed.

The primary analysis was a non-inferiority comparison of the AI-assisted summary against the manual summary on the two co-primary composites, content and presentation, with a non-inferiority margin m = 0.5 composite points tested one-sided at α = 0.025; non-inferiority was concluded for a composite where the lower bound of the two-sided 95% interval for Δ = mean (AI-assisted − manual) lay above −m. Both co-primary endpoints had to succeed (intersection– union), with no multiplicity adjustment across the two co-primary tests. Because the 0.5 margin is pragmatic rather than empirically validated [15], the posterior of Δ is also reported against a stricter 0.25-point benchmark. The primary estimator was a Bayesian hierarchical model with a Gaussian likelihood on the composite, fixed effects for arm and round, and crossed random effects for case, rater and case-by-arm, fitted by the No-U-Turn sampler [16] with weakly informative priors; convergence was required at 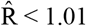 with zero divergent transitions. Prior scales, sampler settings and the full model specification are given in the companion methods preprint [10] and its statistical analysis plan.

Duplicate scoring of each case identifies the between-rater variance component separately from the case-level arm contrast, which the hierarchical model estimates jointly. A frequentist linear mixed model on the within-(case × rater) paired differences, fitted with case as a random effect, rater as a fixed effect, and no case-by-arm interaction, was the pre-specified confirmation analysis (termed the convergence check in the statistical analysis plan), so the two models differ in structure as well as in estimation paradigm. The primary conclusion stood only where the two agreed in direction, and prior robustness was verified by widening the prior scales by factors of 1.0, 2.5, and 5.0.

Secondary analyses were performed to explore performance at the individual-dimension level and to evaluate potential sources of systematic bias. These pre-specified analyses comprised dimension-level non-inferiority on each of the six QSI dimensions, analysed with the same Bayesian model family as the composite, exploratory two-sided superiority tests on each composite, the round fixed effect as a counterbalancing check, and per-rater arm means as a check that the contrast is not driven by a single rater. Inter-rater reliability analyses were performed to quantify the consistency of scoring across evaluators, and are reported as Krippendorff’s α (an ordinal difference function for the raw 1−5 dimensions and an interval function for the continuous composites) [17], with pairwise quadratic-weighted Cohen’s κ [18], Spearman ρ, and exact-agreement percentage for overlapping rater pairs. Power analyses were conducted during study planning to evaluate whether the proposed sample size was sufficient for the primary non-inferiority objective.

The feasibility-determined sample of 38 analysis cases (76 paired observations per endpoint) was confirmed by a design-stage Monte-Carlo power analysis to have ≥0.99 power at the 0.5 margin, assuming a paired-difference standard deviation ≤0.8 and an intra-case correlation ≤0.5; the same analysis attained ≥0.85 power against the stricter 0.25 benchmark only where the paired-difference standard deviation was ≤0.6, so the 0.25 reading is pre-specified as a benchmark rather than as a powered test. The power analysis is not recomputed on observed data, and the observed variance components instead inform the Phase 2 sample-size justification. Multiple-testing adjustments were applied to control the risk of false-positive findings across the dimension-level analyses. Holm adjustment across the six dimensions [19] applies to the frequentist per-dimension tests; on the Bayesian basis no multiplicity adjustment is applied and the unadjusted posterior threshold of 0.975 is retained. The dimension-level analyses should be interpreted with greater caution than the composite analyses because they are based on individual ordinal items rather than aggregated scores. Each dimension is a single 1−5 ordinal item carried on the same continuous-scale approximation used for the six-item composite, which is a heavier approximation for one item than for the mean of six.

## Results

### Cases, analysis sets, and rater coverage

Thirty-seven of 38 planned cases entered the analysis; one case was excluded for a source-document error, which removed both of its arms and left 74 of the 76 planned paired observations. Each analysis case received two independent ratings: one rater scored all 37 cases and two further raters scored complementary subsets of 19 and 18 cases (Figure 1). Second-round cases were not available to a rater until at least fourteen days after that rater’s confirmed completion of the first round; the confirmed completion and assignment dates for each rater are held in the study record. No device deficiencies attributable to the system under evaluation were identified during the study: the observed count was zero in each of the four CPSP severity categories across the 38 designed analysis cases, giving an observed proportion of 0% with an exact (Clopper–Pearson) 95% confidence interval of 0% to 9.3% for each category. The denominator is the 38 designed analysis cases and not the 37 in the complete-case analysis set, since device-deficiency surveillance covered every case processed. The single case exclusion was not a device deficiency and not a processing or export failure of the system under evaluation.

**Figure 1.**
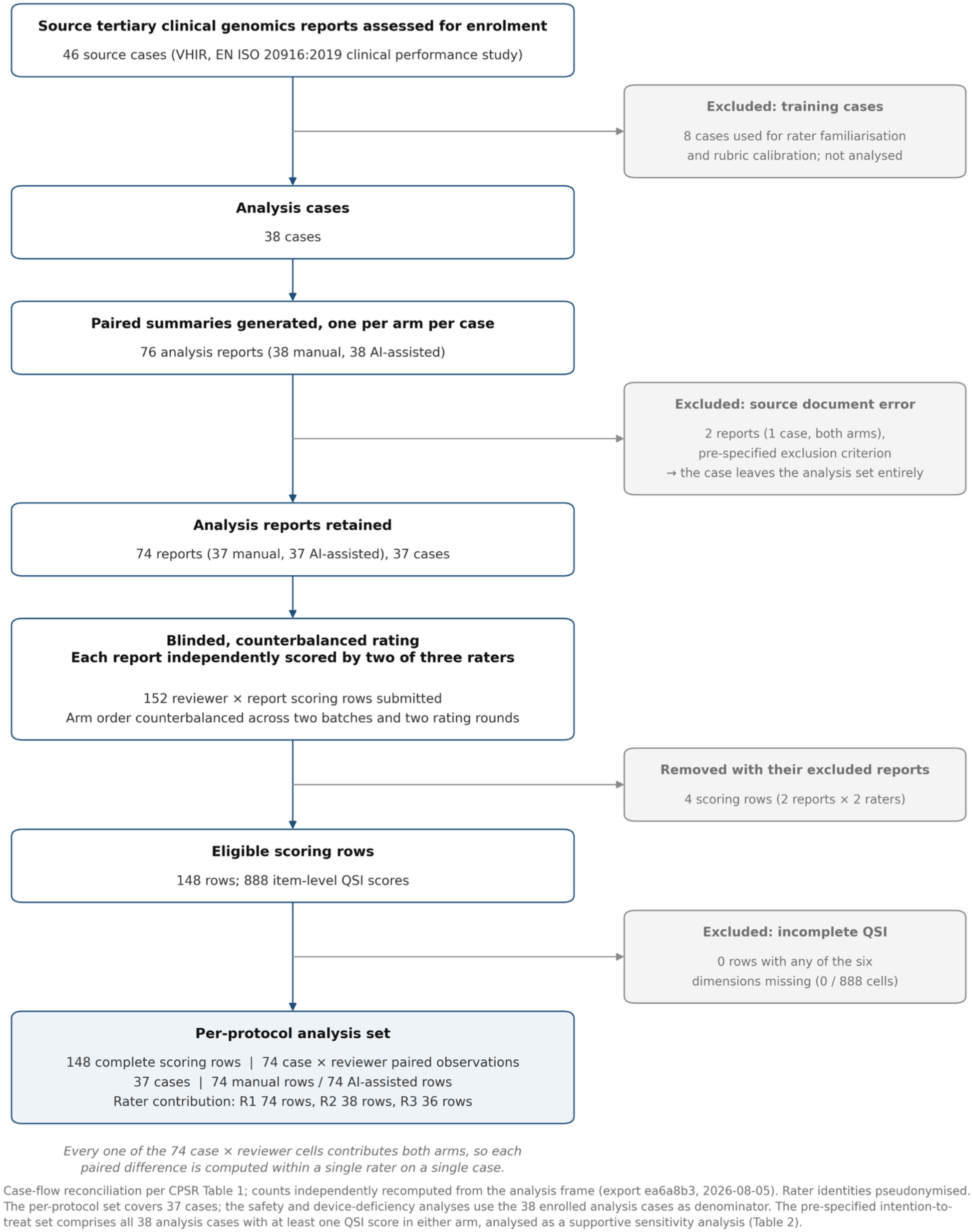
Study-flow diagram (STROBE-style) [20]

### Descriptive QSI scores

In the analysis set (74 paired observations), mean scores are summarised in Table 1. Content composite scores were similar between the AI-assisted and manual arms (4.01 vs 3.97), whereas presentation scores were higher for the AI-assisted arm (4.39 vs 4.12); overall composite scores were also higher for the AI-assisted arm (4.20 vs 4.05). At the dimension level, the largest positive difference was succinctness (+0.69) and the largest negative difference was accuracy (−0.18). The paired within-(case × rater) differences (AI-assisted − manual) had standard deviations of 0.65, 0.74 and 0.57 for content, presentation and overall.

### Primary analysis: co-primary non-inferiority

The Bayesian hierarchical model demonstrated non-inferiority for both co-primary endpoints (Figure 2). Content scores were comparable between the AI-assisted and manual arms, whereas presentation scores were higher for AI-generated summaries; the findings held under the more stringent 0.25-point benchmark. The posterior mean difference (AI-assisted − manual) was +0.05 for the content composite (95% CrI −0.18 to +0.28) and +0.28 for the presentation composite (95% CrI +0.10 to +0.47). At the 0.5 margin, P(Δ > −0.5) > 0.999 for both composites; at the 0.25 benchmark, P(Δ > −0.25) = 0.995 for content and > 0.999 for presentation. The intersection–union condition was satisfied at the 0.5 margin and also at the 0.25 benchmark. Both models converged (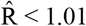 < 1.01, zero divergent transitions).

**Figure 2.**
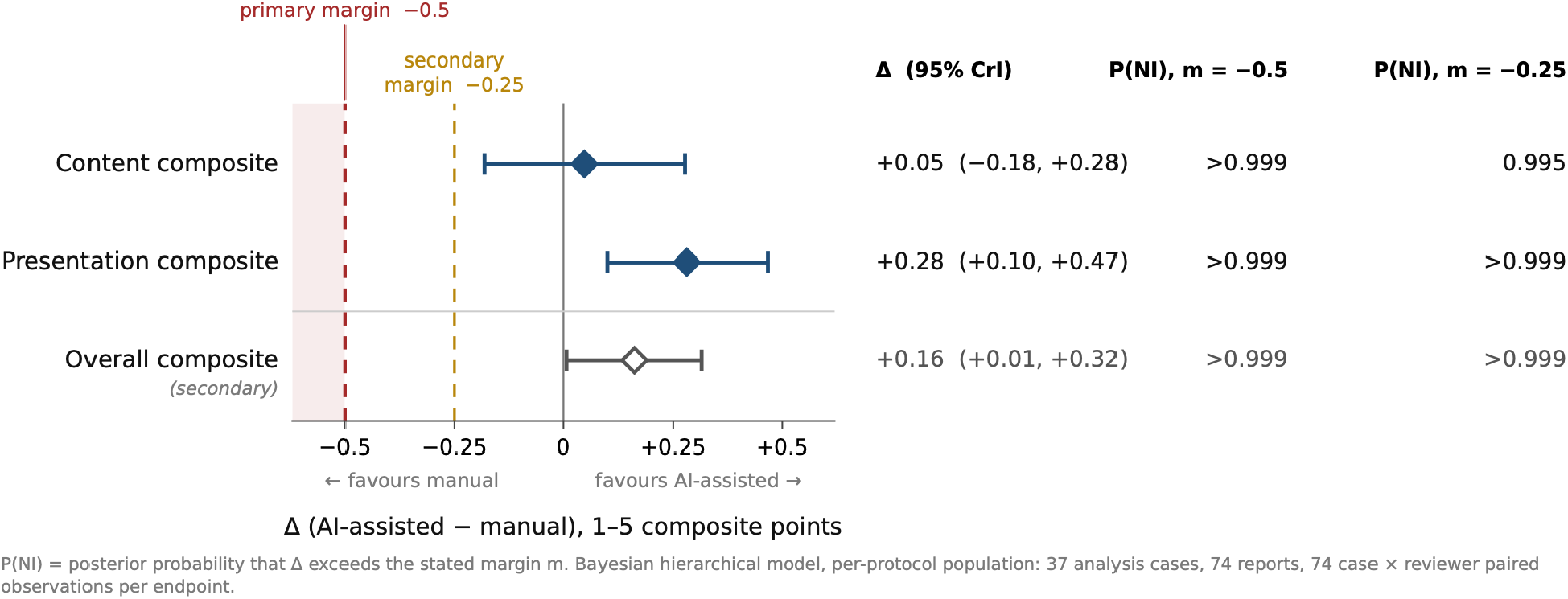
Primary non-inferiority analysis: Δ (95% CrI) for content, presentation and overall composites.

**Figure 3.**
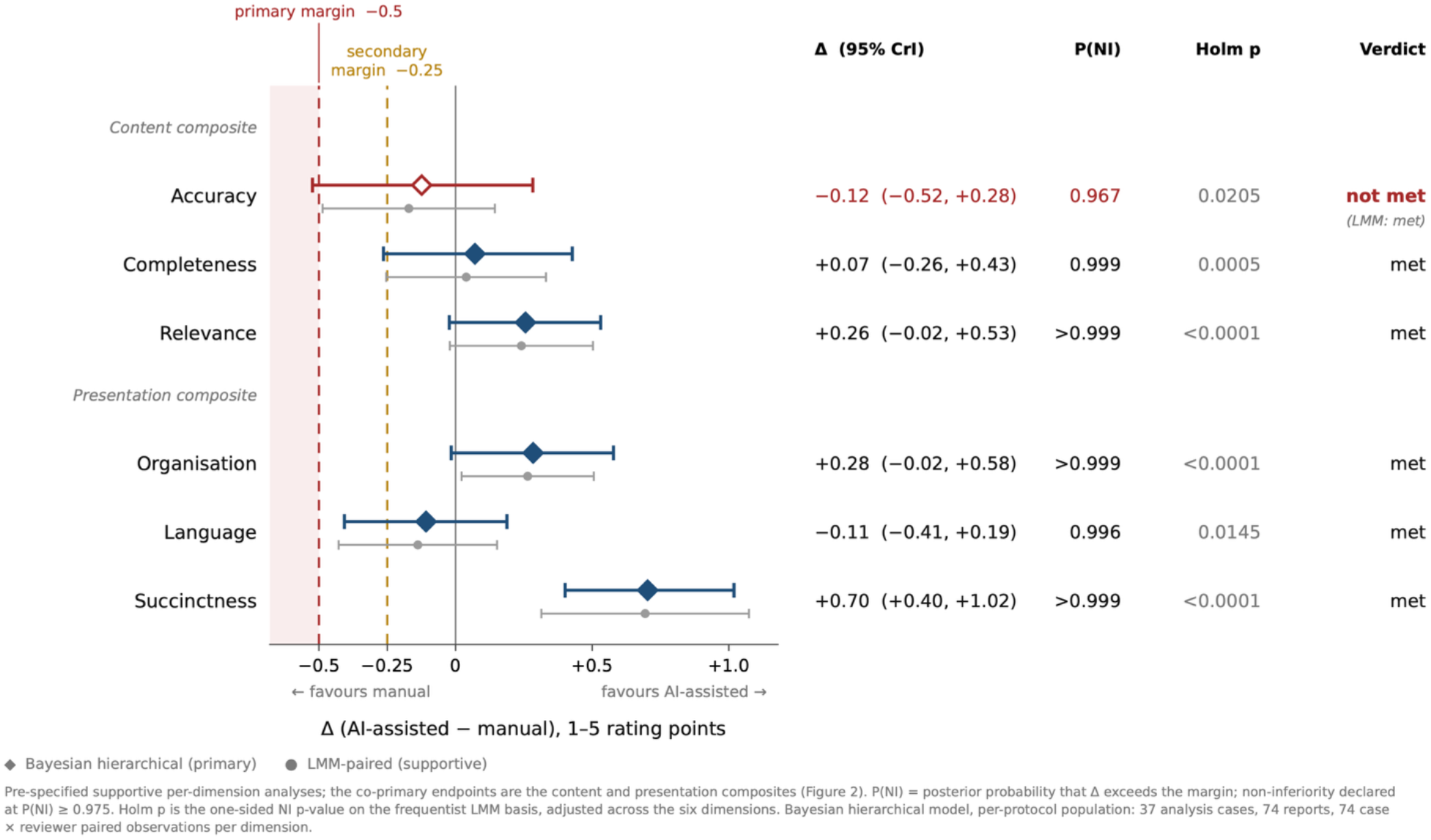
Dimension-level non-inferiority: Δ (95% CrI) for individual QSI dimensions, grouped by composite

### Frequentist confirmation analysis

The pre-specified frequentist linear mixed model on the within-(case × rater) paired differences gave Δ = +0.04 for content (95% CI −0.12 to +0.20) and Δ = +0.27 for presentation (95% CI +0.09 to +0.46); the overall composite gave Δ = +0.16 (95% CI +0.01 to +0.30). Both co-primary estimates agreed in direction and in non-inferiority verdict with the Bayesian primary.

Pre-specified sensitivity analyses, including intention-to-treat and outlier-exclusion analyses, were concordant with the primary findings and did not alter the non-inferiority conclusions (Table 2). All three excluded outlier pairs originated with the same half-roster rater, consistent with the rater heterogeneity reported below. There was no item-level missingness (0 of 888 item scores), so the missing-data refit was not triggered.

**Table 2.** (Results). Concordance of the primary, confirmatory and pre-specified sensitivity analyses on the two co-primary composites.

| Analysis | Content $\Delta$ (95% interval) | Presentation $\Delta$ (95% interval) | Conclusion |
| --- | --- | --- | --- |
| Bayesian primary (per-protocol) | +0.05 (−0.18 to +0.28) | +0.28 (+0.10 to +0.47) | Non-inferior |
| Frequentist confirmation (LMM) | +0.04 (−0.12 to +0.20) | +0.27 (+0.09 to +0.46) | Non-inferior |
| Intention-to-treat sensitivity | +0.04 (−0.19 to +0.26) | +0.29 (+0.10 to +0.47) | Non-inferior |
| Outlier exclusion (>3 SD) | +0.11 (−0.11 to +0.32) | +0.25 (+0.06 to +0.43) | Non-inferior |
| Prior sensitivity (widest scale)* | +0.04 | +0.27 | Non-inferior (diagnostic) |

\*Bayesian intervals are 95% credible intervals; the frequentist row gives a 95% confidence interval. Prior-sensitivity refits at the widest scale did not meet the convergence criterion set for the primary fits and are reported as diagnostics only (Table 3).

### Prior-robustness

Because Bayesian models require specification of prior assumptions, sensitivity analyses were conducted to confirm that the study conclusions were not dependent on the chosen priors. To further assess the robustness of the Bayesian findings, prior scales were progressively widened by factors of 1.0, 2.5 and 5.0 and the resulting estimates compared with the primary analysis. Both composites were classified prior-robust, with P(Δ > −0.5) > 0.999 at every scale (Table 3). At the widest scale neither refit met the convergence criterion set for the primary fits, so those refits are reported as diagnostics; the primary fits met the criterion and no reported verdict depends on them.

**Table 3.** (Results). Prior-sensitivity analysis: posterior mean difference and non-inferiority conclusion for each co-primary composite at each pre-specified prior scale.

| Prior scale | Composite | $\Delta$ (95% CrI) | $P(\Delta > -0.5)$ | Max $\hat{R}$ | Divergent transitions |
| --- | --- | --- | --- | --- | --- |
| 1.0 (primary) | Content | +0.05 (−0.18 to +0.28) | >0.999 | 1.000 | 0 |
| 1.0 (primary) | Presentation | +0.28 (+0.10 to +0.47) | >0.999 | 1.000 | 0 |
| 2.5 | Content | +0.04 (−0.19 to +0.26) | >0.999 | 1.000 | 0 |
| 2.5 | Presentation | +0.28 (+0.08 to +0.47) | >0.999 | 1.000 | 0 |
| 5.0 (widest) | Content | +0.04 (−0.18 to +0.25) | >0.999 | **1.010** | 0 |
| 5.0 (widest) | Presentation | +0.27 (+0.08 to +0.47) | >0.999 | 1.000 | **6** |

$\Delta$ is the posterior mean difference (AI-assisted – manual) on the 1–5 composite scale; intervals are 95% credible intervals. $\hat{R}$ is the maximum across model parameters. The convergence criterion set for the primary fits is $\hat{R} < 1.01$ with zero divergent transitions; the two refits at the widest scale fell outside it, on different criteria, and are reported as diagnostics only. Non-inferiority was concluded at the 0.5 margin for all six refits.

### Secondary: dimension-level non-inferiority

Secondary analyses explored performance across individual QSI dimensions and identified potential sources of difference masked by the composite endpoints. The per-dimension secondary applies the composite Bayesian model family to each dimension; Holm adjustment across the six [19] applies to the frequentist tests reported alongside. Five of the six dimensions met non-inferiority at the 0.5 margin: completeness (posterior mean +0.07, P(Δ > −0.5) = 0.999), relevance (+0.26, > 0.999), organisation (+0.28, > 0.999), language (−0.11, 0.996), and succinctness (+0.70, > 0.999).

Accuracy was the exception: posterior mean −0.12 (95% CrI −0.52 to +0.28), P(Δ > −0.5) = 0.967, below the 0.975 decision threshold, so non-inferiority was not concluded for that dimension. The frequentist linear mixed model with Holm adjustment reached non-inferiority on all six dimensions including accuracy (adjusted p = 0.0205), so the two models disagree on accuracy alone. The frequentist model treats rater as a fixed effect and omits the case-by-arm term, so a disagreement is expected to surface first on the dimension carrying the largest between-rater component, as we observed for accuracy. The per-dimension estimates, with the frequentist results alongside, are tabulated in the Supplementary Information (S8).

### Exploratory: superiority

Exploratory superiority analyses were performed to evaluate whether any domains favoured the AI-assisted arm beyond the non-inferiority framework. In pre-labelled exploratory two-sided superiority tests, the presentation composite showed a superiority signal (P(Δ > 0) = 0.999) as did the overall composite (Δ = +0.16, 95% CrI +0.01 to +0.32; P(Δ > 0) = 0.979); the content composite did not (P(Δ > 0) = 0.663). At the dimension level succinctness favoured the AI-assisted arm on both estimators (Bayesian P(Δ > 0) > 0.999); organisation reached the superiority threshold on the frequentist model (Δ = +0.26, 95% CI +0.02 to +0.51) but not on the Bayesian (P(Δ > 0) = 0.968), and relevance fell just short on both (Bayesian P(Δ > 0) = 0.964). These tests are exploratory and do not displace the non-inferiority framing.

### Counterbalancing and rater checks

Counterbalancing and rater checks were performed to assess whether the observed differences between study arms could be explained by period effects or by the scoring behaviour of individual raters. The round fixed effect was consistent with zero for every composite (content +0.01, 95% CrI −0.20 to +0.22; presentation −0.10, 95% CrI −0.29 to +0.09; overall −0.05, 95% CrI −0.21 to +0.11), so no material period effect confounded the arm contrast. The direction of the arm contrast was also consistent across raters: on the overall composite each of the three raters scored the AI-assisted arm at or above the manual arm (per-rater AI-assisted − manual differences of +0.21, +0.17, and +0.04), so the contrast is not attributable to a single rater.

### Inter-rater reliability

Inter-rater reliability analyses were performed to assess the consistency of scoring across evaluators. Agreement was low across most QSI dimensions and composites, and one rater scored consistently higher than the other two. Krippendorff’s α [17] (ordinal function for the six raw dimensions, interval function for the composites) was close to zero or negative for most elements, and weighted Cohen’s κ [18], Spearman ρ and exact-agreement rates followed the same pattern (Table 4). Succinctness and relevance reached the highest pooled agreement, and the arm-stratified estimates were not symmetric across arms: succinctness agreement was carried by the manual arm and language agreement by the AI-assisted arm, while the presentation composite reached its highest value in the AI-assisted arm alone (α = +0.17 against −0.06 manual). For the remaining dimensions and composites the arm-stratified values were near zero or negative in both arms (Supplementary Information, S1). Cohen’s κ was computable only for the two overlapping rater pairs, each involving the all-cases rater and co-rating 36 and 38 reports; the two half-roster raters scored disjoint subsets and never co-rated a case. Scores therefore depended substantially on which rater assigned them, which bears on the dimension-level results. ICC(2,1) is not reported because the rater roster is unbalanced [21].

**Table 4.**
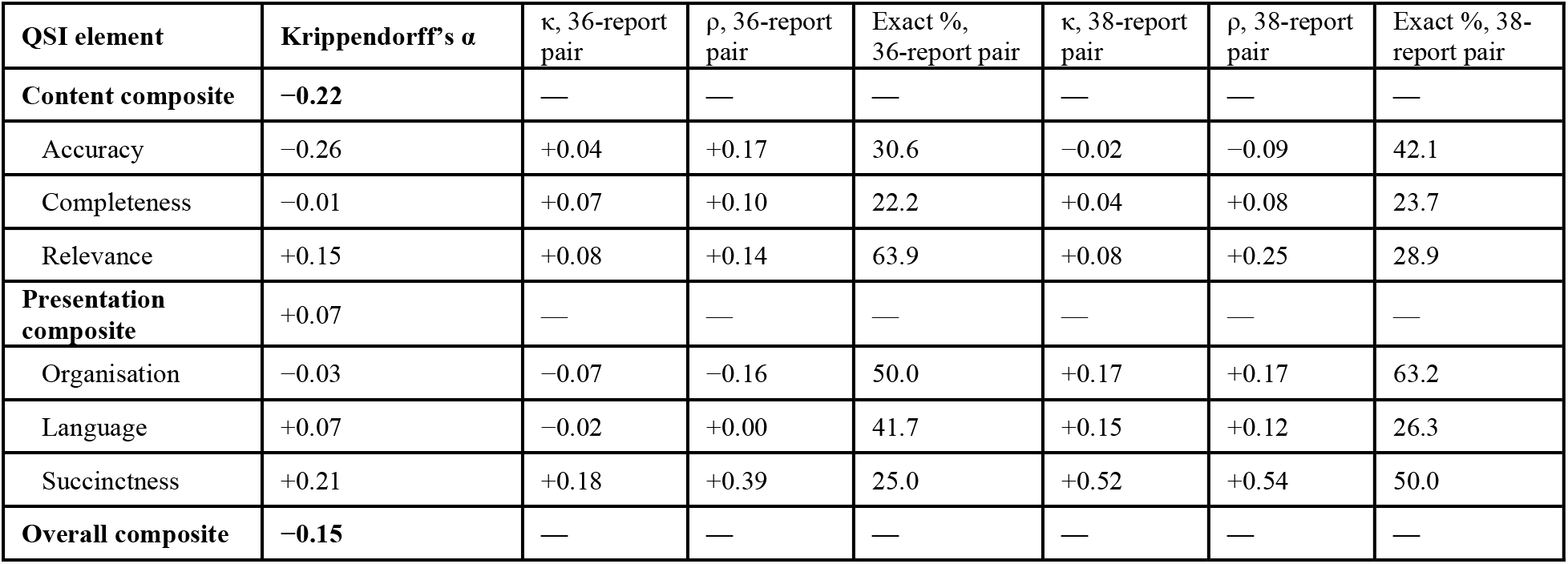
(Results). Inter-rater agreement by QSI element. Cohen’s κ, Spearman ρ and exact agreement are computed for the two overlapping rater pairs on the six raw dimensions and are not defined for the composites.

Krippendorff’s α is reported as point estimates; interval estimation was not pre-specified and bootstrap confidence intervals were not computed, so the α values are descriptive of this roster and carry no quantified precision.

## Discussion

This retrospective blinded study compared AI-generated and manually generated summaries of molecular reports, with both summary types evaluated against the same source report. The primary finding was that AI-generated summaries preserved clinically relevant content at a level comparable to manual summaries while achieving higher presentation quality. Accuracy was the weakest-performing dimension and the only one for which the Bayesian and frequentist analyses yielded differing conclusions.

Clinical text summarisation by large language models has been compared with clinician-written summaries in several settings, and adapted models have been reported to match or exceed them [7]. The instruments used to make those judgements are less well established [3,4,5], and where a validated instrument has been used it has been applied to general clinical documentation: the PDSQI-9 was validated on electronic health record notes, where physician raters reached an intraclass correlation coefficient of 0.867, and it has since served as the human reference for automated evaluation of clinical summaries [13,22]. This study applies that approach to tertiary genomics reports, under endpoints and an analysis plan published before data lock [10,11], and tests non-inferiority against a manual comparator drawn from the same source report. The QSI did not reach the agreement its parent instrument achieved on clinical notes, so the present limit on evaluating genomic-report summarisation lies in the instrument as applied by these raters.

The asymmetry between the two co-primary composites was anticipated in the analysis plan, with non-inferiority expected on content and possible superiority reserved for presentation and succinctness. Both arms summarise the same source molecular report, and the accuracy dimension scores concordance with that source, so a comparator summary prepared under the standard VHIR workflow by the professionals who issue and interpret these reports leaves limited scope for a higher fidelity score, while structure and length were open to improvement in either arm. The observations and exploratory superiority tests are consistent with those expectations, and succinctness showed the largest dimension-level difference. A ceiling on the content composite does not, however, account for a point estimate favouring the manual arm on accuracy, which was also the dimension with the lowest inter-rater agreement, the widest posterior, and the largest score dispersion of the six.

Low inter-rater reliability was anticipated for a pragmatic three-rater roster in which one rater scores every case, and the posterior variance components place the dominant source of variation with the raters, with the case and arm-by-case components smaller (Supplementary Information, S2). The QSI, as worded and as applied by these raters to these reports, did not produce a shared standard across raters; accuracy, with the lowest agreement and the largest dispersion of the six dimensions, is where that is most visible, and it is also the dimension on which the two analysis models disagreed. The non-inferiority verdict therefore rests on a hierarchical model that estimates the case-level arm contrast while regularising a large between-rater variance component, so the instrument separated summaries less sharply than the composite point estimates alone suggest. Because the rater standard deviation is estimated from three raters and is weakly identified, any Phase 2 sample-size determination that rests on it should be treated as provisional.

As clinicians who read tertiary genomics reports in practice, we note that the qualities the QSI measures are the attributes that reporting guidelines ask of the source reports themselves: European and ESMO recommendations both require that a genomic report answer the clinical question clearly and concisely, and that it be written so that a physician who is not a genomicist can act on the result [23,24]. The higher presentation scores of the AI-assisted summaries are therefore relevant to that goal: readability, organisation and succinctness may facilitate the communication of complex genomic findings to clinicians with varying levels of genomics expertise. In day-to-day practice, readability, succinctness, and reporting in the clinician’s local language reduce cognitive burden. Application of the six QSI dimensions was feasible across the reports evaluated, although some dimensions appeared operationally overlapping. The distinction between factual concordance with the source report (Accuracy) and clinical usefulness of the retained information (Relevance) was not always straightforward in practice and may have contributed to inter-rater variability. In particular, assessing Relevance often required substantial expert judgement because its interpretation depended on the clinical context of the individual case. Similarly, distinguishing between Relevance and Succinctness, or between Accuracy and Completeness, sometimes required deliberation, as a summary may faithfully reflect the source report while differing in the amount and clinical usefulness of the information retained. Some of the inter-rater variability may therefore reflect the difficulty of operationalising partially overlapping dimensions in the current QSI, in addition to differences between raters. Across both approaches, summaries were generally well organised, readable and faithful to the underlying genomic findings. Common limitations included variability in the level of detail provided, limited incorporation of patient-specific clinical context, and trade-offs between comprehensiveness and conciseness. Although these observations were not collected systematically and should be considered qualitative, they complement the reliability findings and may help guide future refinement of the QSI definitions, scoring anchors and rater calibration prior to prospective evaluation.

Several features of the design strengthen these findings. The endpoints and the analysis were pre-specified and published before data lock. The Bayesian primary analysis was assessed against a frequentist model and prior-sensitivity analyses, and the primary conclusion was required to survive both. Blinding was implemented at the case and arm level, with arm counterbalanced against round and a washout enforced between a rater’s two scoring events for the same case. The rubric is anchored, its anchors are published in the companion preprint, and rater agreement is reported. The analysis was executed by sponsor personnel against the published plan and frozen before unblinding, with the analysis code held under version control and validated by independent reading; all reported results derive from a single locked export.

Several limitations qualify these findings. The reliability results are conditional on the single all-cases rater, and the analysis plan adopted a Bayesian primary because a three-rater frequentist estimator of the rater variance would be near-singular. The QSI’s content validity for the specific failure modes of tertiary genomics summarisation has not been independently established. This is a single site, a single curated paediatric oncology dataset, and one system under evaluation, so the design describes a scoring instrument applied to a single tool rather than a comparison across tools, and rubric behaviour may not generalise to other centres or to adult and non-oncology genomics. The sample size was feasibility-determined, so a result short of the threshold would represent absence of demonstrated non-inferiority rather than evidence of equivalence [25]. The composite is treated as continuous under a Gaussian likelihood as a tractability choice, and the per-dimension secondary carries the same approximation on single 1–5 ordinal items, where it is heavier and where the one undemonstrated verdict arose. Blinding integrity was supported procedurally but not formally assessed; and the 0.5 margin is pragmatic, which is why conclusions are also read against the 0.25 benchmark: because the design-stage Monte-Carlo grid gives roughly 0.66 power for content and 0.78 for presentation against a 0.25 margin at the model-implied paired-difference standard deviations (0.94 and 0.80) when read from that grid without recomputation (Supplementary Information, S4), the 0.25 result is a supportive benchmark and not a powered confirmatory test. Finally, the assessors are co-authors of this report; the primary measurement preceded any authorship role and was made under blinding to summary origin, which reduces but does not remove the potential for assessor expectation to influence scores in a sponsor-funded evaluation. The study evaluates summary quality under retrospective blinded assessment and therefore does not provide evidence regarding clinical utility, workflow integration, user adoption, or effects on clinical decision-making. These questions remain for future prospective evaluation.

The dominant source of variability was the raters rather than the cases, so QSI refinement and rater calibration precede any larger prospective study. The posterior variance components, the realised paired-difference standard deviations and the implied intra-case correlations that inform Phase 2 sample-size planning are reported in the Supplementary Information (S2, S4 and S5); the rater standard deviation is estimated from three raters and should be reassessed after QSI-anchor refinement and rater calibration before it informs that planning. Consistent with DECIDE-AI recommendations [26], clinical utility, workflow integration and decision-support performance were outside the scope of this retrospective study.

In this retrospective blinded evaluation, AI-generated summaries were non-inferior to manually generated summaries on content and and scored higher on presentation. Accuracy was not shown non-inferior and inter-rater agreement was low, so the instrument and the rater calibration are priority for the next phase. Clinical validity and workflow effect were not assessed, and these findings support further prospective evaluation of AI-assisted genomic-report summarisation in routine practice.

## Declarations

### Ethics approval and consent

Governed by the CEIM at VHIR; favourable opinion including a waiver of additional informed consent for the non-interventional use of de-identified archived data under reference PR(AMI)318-2025, dated 08 August 2025. Conducted in accordance with the Declaration of Helsinki [27] and applicable data-protection law, including the EU GDPR [28] and the Swiss FADP [29]; de-identification, the Data Protection Impact Assessment, and the legal basis for data transfer are specified in the study data-protection documentation.

### Data availability

The QSI rubric and the randomisation and counterbalancing procedure are available from the corresponding author on reasonable request. All results reported here derive from a single locked analysis export (analysis code commit ea6a8b3, exported 05 August 2026) comprising 152 raw scoring rows, of which 148 entered the complete-case analysis set. De-identified source reports and individual rater scores are governed by the CEIM approval and applicable data-protection law and are held under controlled access at VHIR with a ten-year retention period; they are not publicly available. Supporting statistical output is provided in the Supplementary Information (Supplementary Statistical Validation sections S1 to S8); the pre-specified statistical analysis plan is published with the companion methods preprint [10].

Code availability. The AI-assisted summarisation system under evaluation is a commercial product and its source code is not available. The analysis code implementing the pre-specified statistical analysis plan is available to the editors and referees on request; the plan itself, including the model specifications, priors and convergence criteria, is published in full in the companion methods preprint [10]. Analyses were performed in Python 3.13 using NumPyro 0.21.0 for the Bayesian primary analysis and statsmodels 0.14.6 for the frequentist confirmation analysis.

### Author contributions (CRediT)

J.C. Conceptualization, Methodology, Investigation, Writing original draft; M.O. Data curation, Formal analysis, Software, Validation, Methodology; A.S. Conceptualization, Methodology, Resources, Supervision, Project administration, Writing review and editing. N.B., R.H.A., and L.V.A. Investigation (blinded QSI scoring), Validation (QSI application and calibration), Writing review and editing. A.C.T. Project administration, Resources, and Writing review and editing.

## Acknowledgements

We are particularly grateful to Manuel Laguía for his continued support in facilitating and coordinating the collaboration between VHIR and Qnomx AG throughout the project. We also thank Rafael Navajo and Nunzio G. Cifariello for their support in establishing this collaboration, and Miguel F. Segura and Joerg Hoelzing for their roles in initiating and fostering this partnership.

## References

1. Gibson JS, El Achi H, Altenburger D, et al. Developing consensus for a more provider-friendly next-generation sequencing molecular biomarker report: a joint consensus recommendation of the Association for Molecular Pathology and College of American Pathologists. J Mol Diagn. 2025;27(12):1123–36. doi:10.1016/j.jmoldx.2025.08.011. PMID: 41016627

2. Li MM, Datto M, Duncavage EJ, et al. Standards and guidelines for the interpretation and reporting of sequence variants in cancer: a joint consensus recommendation of the Association for Molecular Pathology, American Society of Clinical Oncology, and College of American Pathologists. J Mol Diagn. 2017;19(1):4–23. doi: 10.1016/j.jmoldx.2016.10.002. PMID: 27993330

3. Tam TYC, Sivarajkumar S, Kapoor S, et al. A framework for human evaluation of large language models in healthcare derived from literature review. npj Digit Med. 2024;7(1):258. doi:10.1038/s41746-024-01258-7. PMID: 39333376

4. Bedi S, Liu Y, Orr-Ewing L, et al. Testing and evaluation of health care applications of large language models: a systematic review. JAMA. 2025;333(4):319–28. doi:10.1001/jama.2024.21700. PMID: 39405325

5. Bednarczyk L, Reichenpfader D, Gaudet-Blavignac C, et al. Scientific evidence for clinical text summarization using large language models: scoping review. J Med Internet Res. 2025;27:e68998. doi:10.2196/68998. PMID: 40371947.

6. Adams G, Zucker J, Elhadad N. A meta-evaluation of faithfulness metrics for long-form hospital-course summarization. Proc Mach Learn Res. 2023;219:2–30.

7. Van Veen D, Van Uden C, Blankemeier L, et al. Adapted large language models can outperform medical experts in clinical text summarization. Nat Med. 2024;30(4):1134–42. doi:10.1038/s41591-024-02855-5. PMID: 38413730

8. Wang J, Li H, Liu H. A comprehensive system for searching and evaluating genomic variant evidence using AI and knowledge bases to support personalized medicine. AMIA Annu Symp Proc. 2025;2024:1206–14. PMID: 40417484.

9. Cannon M, Bratulin A, Kuzma K, et al. Transforming semi-structured variant assessments into computable clinical assertions: a pilot study for AI-assisted curation. medRxiv. 2026. doi:10.64898/2026.05.07.26352456.

10. Creeden J, Olivecrona M, Soriano A. A blinded, counterbalanced rater design for evaluating AI-assisted summarisation of tertiary clinical genomics reports: methodology of the QNOMX-VHIR-CPSP-001 Phase 1 study. medRxiv. 2026. doi:10.64898/2026.06.11.26355467.

11. European Committee for Standardization. In vitro diagnostic medical devices: clinical performance studies using specimens from human subjects: good study practice (ISO 20916:2019). EN ISO 20916:2024. Brussels: CEN; 2024.

12. Vilaplana A, Carolina Torres A, Escudero López A, et al. Establishing a multicentre personalised medicine programme in childhood, adolescent and young adult cancer in Spain: the SEHOP-PENCIL project. JCO Precis Oncol. 2026 Aug;10(9):e2600074. doi: 10.1200/PO-26-00074. Epub 2026 Sep 1. PMID: 42679328

13. Croxford E, Gao Y, Pellegrino N, et al. Development and validation of the Provider Documentation Summarization Quality Instrument for large language models. J Am Med Inform Assoc. 2025;32(6):1050–60. doi:10.1093/jamia/ocaf068. PMID: 40323321

14. Stetson PD, Bakken S, Wrenn JO, Siegler EL. Assessing electronic note quality using the Physician Documentation Quality Instrument (PDQI-9). Appl Clin Inform. 2012;3(2):164–74. doi:10.4338/ACI-2011-11-RA-0070. PMID: 22577483

15. European Medicines Agency, Committee for Proprietary Medicinal Products. Guideline on the choice of the noninferiority margin. EMEA/CPMP/EWP/2158/99. London: EMEA; 2005.

16. Hoffman MD, Gelman A. The No-U-Turn sampler: adaptively setting path lengths in Hamiltonian Monte Carlo. J Mach Learn Res. 2014;15:1593–623.

17. Hayes AF, Krippendorff K. Answering the call for a standard reliability measure for coding data. Commun Methods Meas. 2007;1(1):77–89. doi:10.1080/19312450709336664

18. Cohen J. Weighted kappa: nominal scale agreement with provision for scaled disagreement or partial credit. Psychol Bull. 1968;70(4):213–20. doi:10.1037/h0026256. PMID: 19673146

19. Holm S. A simple sequentially rejective multiple test procedure. Scand J Stat. 1979;6(2):65–70.

20. von Elm E, Altman DG, Egger M, et al.; STROBE Initiative. The Strengthening the Reporting of Observational Studies in Epidemiology (STROBE) statement: guidelines for reporting observational studies. Lancet. 2007;370(9596):1453–7. doi:10.1016/S0140-6736(07)61602-X. PMID: 17941714

21. Koo TK, Li MY. A guideline of selecting and reporting intraclass correlation coefficients for reliability research. J Chiropr Med. 2016;15(2):155–63. doi:10.1016/j.jcm.2016.02.012 PMID: 27330520

22. Croxford E, Gao Y, First E, et al. Evaluating clinical AI summaries with large language models as judges. npj Digit Med. 2025;8:640. doi:10.1038/s41746-025-02005-2.

23. Deans ZC, Ahn JW, Carreira IM, et al. Recommendations for reporting results of diagnostic genomic testing. Eur J Hum Genet. 2022;30(9):1011–6. doi:10.1038/s41431-022-01091-0. PMID: 35572999

24. van de Haar J, Roepman P, Andre F, et al. ESMO recommendations on clinical reporting of genomic test results for solid cancers. Ann Oncol. 2024;35(11):954–67. doi:10.1016/j.annonc.2024.06.018. PMID: 39112111

25. Piaggio G, Elbourne DR, Pocock SJ, Evans SJW, Altman DG; CONSORT Group. Reporting of noninferiority and equivalence randomized trials: extension of the CONSORT 2010 statement. JAMA. 2012;308(24):2594–604. doi:10.1001/jama.2012.87802. PMID: 23268518

26. Vasey B, Nagendran M, Campbell B, et al. Reporting guideline for the early-stage clinical evaluation of decision support systems driven by artificial intelligence: DECIDE-AI. Nat Med. 2022;28(5):924–33. doi:10.1038/s41591-022-01772-9. PMID: 35585198

27. World Medical Association. World Medical Association Declaration of Helsinki: ethical principles for medical research involving human participants. JAMA. 2025;333(1):71–4. doi:10.1001/jama.2024.21972. PMID: 39425955

28. Regulation (EU) 2016/679 of the European Parliament and of the Council of 27 April 2016 (General Data Protection Regulation). Off J Eur Union. 2016;L119:1–88.

29. Switzerland. Federal Act on Data Protection (FADP) of 25 September 2020, SR 235.1. In force 1 September 2023.

