## Supplementary Information for "Evaluation of AI-assisted summarisation of tertiary clinical genomics reports: results of the QNOMX-VHIR-CPSP-001 Phase 1 study"

All values derive from a single locked analysis export of the QNOMX-VHIR-CPSP-001 Phase 1 dataset (analysis code commit ea6a8b3, exported 05 August 2026). The statistical analysis plan, including the prior specifications, sampler settings, convergence criterion, analysis-set definitions and multiplicity treatment, is published in the companion methods preprint [10].

“AI-assisted” denotes summaries produced by the AI-assisted summarisation system under evaluation and “manual” denotes summaries produced by the standard workflow. Δ is the difference (AI-assisted − manual) on the 1–5 composite or dimension scale.

### S1. Arm-stratified inter-rater agreement

Table 4 of the main text reports Krippendorff’s α pooled across arms. Table S1 gives the arm-stratified estimates, using the ordinal difference function for the six raw QSI dimensions and the interval function for the three composites [17].

**Table S1.** Krippendorff’s α by QSI element and arm. Pooled n = 74 scoring units; each arm n = 37. The manual-arm completeness estimate is −0.0005.

| QSI element | Pooled α | AI-assisted arm α | Manual arm α |
|---|---|---|---|
| **Content composite** | −0.22 | −0.31 | −0.08 |
| Accuracy | −0.26 | −0.28 | −0.24 |
| Completeness | −0.01 | −0.02 | 0.00 |
| Relevance | +0.15 | +0.16 | +0.11 |
| **Presentation composite** | +0.07 | +0.17 | −0.06 |
| Organisation | −0.03 | −0.04 | −0.06 |
| Language | +0.07 | +0.33 | −0.20 |
| Succinctness | +0.21 | −0.04 | +0.31 |
| **Overall composite** | −0.15 | −0.26 | −0.07 |

Agreement was arm-dependent for language and succinctness, each positive in one arm and negative in the other.

Interval estimation for α was not pre-specified. The arm-stratified estimates are computed at n = 37 per arm and are correspondingly less precise than the pooled values in Table 4 of the main text.

### S2. Posterior variance components

The primary Bayesian hierarchical model carries a residual term ($\sigma$), crossed random effects for case ($\sigma\_case$) and rater ($\sigma\_rater$), and a case-by-arm interaction ($\sigma\_caseArm$).

**Table S2.** Posterior variance components, implied case-level intraclass correlation, and implied within-(case × rater) paired-difference standard deviation. Posterior means with 95% credible intervals.

| Component | Content | Presentation | Overall |
|---|---|---|---|
| σ (residual) | 0.659 (0.577 to 0.758) | 0.551 (0.476 to 0.637) | 0.467 (0.405 to 0.537) |
| σ_case | 0.449 (0.285 to 0.633) | 0.259 (0.081 to 0.420) | 0.292 (0.168 to 0.424) |
| σ_rater | 1.421 (0.494 to 2.652) | 1.241 (0.235 to 2.674) | 1.341 (0.369 to 2.734) |
| σ_caseArm | 0.119 (0.003 to 0.331) | 0.155 (0.005 to 0.380) | 0.104 (0.004 to 0.290) |
| Implied case-level ICC | 0.318 | 0.181 | 0.281 |
| Implied paired-difference SD (sd_d) | 0.939 | 0.795 | 0.669 |
| Implied intra-case correlation (ρ_case) | 0.016 | 0.038 | 0.024 |

The rater component is the largest source of variation on every composite and the case-by-arm interaction the smallest.

The implied paired-difference SD is derived from the posterior variance components and exceeds the empirical standard deviations of the observed paired differences reported in Table 1 of the main text (0.65, 0.74 and 0.57).

The case-level ICC is the proportion of total score variance attributable to the case. The intra-case correlation $\rho\_case$ is on the within-(case × rater) difference scale and corresponds to the $\rho$ parameter of the design-stage power grid (S4).

### S3. Identifiability of the rater variance component

The rater standard deviation is estimated from three raters, and its 95% credible intervals span roughly an order of magnitude on all three composites (Table S2). The posterior mean is weakly identified, and any inference resting on its magnitude is only indicative.

The analysis plan adopted a Bayesian primary estimator on these grounds, since a frequentist estimator of a rater variance component from three levels is near-singular. The pre-specified frequentist model treats rater as a fixed effect and omits the case-by-arm term, so the two models differ in structure as well as in estimation paradigm, and a disagreement between them is expected to appear first on the dimension carrying the largest between-rater component, as it did on accuracy (S8).

The non-inferiority verdict rests on the case-level arm contrast, which is estimated within case-by-rater cells and does not take the rater main effect as an input. The prior-sensitivity sweep (S6) shows the co-primary verdicts to be stable across a fivefold widening of the prior scales.

Phase 2 sample-size determination resting on σ_rater is provisional and should be recomputed after QSI-anchor refinement and rater calibration (S5).

### S4. Design-stage Monte-Carlo power analysis

The pre-specified power statement is that 38 analysis cases scored across two arms by one all-cases rater plus one of two half-roster raters (76 paired observations per endpoint), assuming truly equivalent arms with a paired-difference standard deviation ≤ 0.8 and intra-case correlation ≤ 0.5, gives ≥ 0.99 power to declare non-inferiority at the 0.5-point margin (one-sided $\alpha = 0.025$) on each co-primary composite. Non-inferiority at the stricter 0.25-point margin is powered (≥ 0.85) only where the paired-difference standard deviation is ≤ 0.6.

**Table S4a.** Design-stage power, arms truly equivalent ($\Delta = 0$).

| Margin | sd_d | ρ = 0.0 | ρ = 0.3 | ρ = 0.5 |
|---|---|---|---|---|
| 0.25 | 0.4 | 0.9998 | 0.9975 | 0.9935 |
| 0.25 | 0.6 | 0.9523 | 0.8873 | 0.8370 |
| 0.25 | 0.8 | 0.7783 | 0.6678 | 0.6218 |
| 0.25 | 1.0 | 0.6080 | 0.4830 | 0.4335 |
| 0.50 | 0.4 | 1.0000 | 1.0000 | 1.0000 |
| 0.50 | 0.6 | 1.0000 | 1.0000 | 1.0000 |
| 0.50 | 0.8 | 0.9988 | 0.9973 | 0.9908 |
| 0.50 | 1.0 | 0.9873 | 0.9698 | 0.9445 |

**Table S4b.** Design-stage power, AI-assisted arm slightly worse ($\Delta = -0.10$), $\rho = 0.3$.

| Margin | sd_d | Power |
|---|---|---|
| 0.25 | 0.4 | 0.8178 |
| 0.25 | 0.6 | 0.4900 |
| 0.25 | 0.8 | 0.3245 |
| 0.25 | 1.0 | 0.2165 |
| 0.50 | 0.4 | 1.0000 |
| 0.50 | 0.6 | 0.9988 |
| 0.50 | 0.8 | 0.9635 |
| 0.50 | 1.0 | 0.8635 |

#### S4.1 Interpolation of the power grid at the model-implied dispersion

At the model-implied paired-difference standard deviations of 0.939 for content and 0.795 for presentation (S2), neither of which is a tabulated grid point, power at the 0.25 margin is interpolated linearly between the bracketing rows of Table S4a:

- Content, sd_d = 0.939, between sd_d = 0.8 (0.7783) and sd_d = 1.0 (0.6080) at the $\rho$ = 0.0 column: $0.7783 - 0.695 \times (0.7783 - 0.6080) = 0.660$.
- Presentation, sd_d = 0.795, between sd_d = 0.6 (0.9523) and sd_d = 0.8 (0.7783).
- Overall composite, sd_d = 0.669: 0.892.

The $\rho = 0.0$ column is used because the model-implied intra-case correlations on the difference scale are 0.016 and 0.038 (S2). Interpolating the $\rho$ axis as well, between the $\rho = 0.0$ and $\rho = 0.3$ columns at those values, gives 0.654 and 0.769, so the choice of column is immaterial at these correlations.

These figures interpolate the design-stage grid at variance components estimated after data lock, and the 0.25 margin is pre-specified as a supportive benchmark. Power at the 0.5 margin exceeds 0.94 at every cell of Table S4a, including sd_d = 1.0 at $\rho = 0.5$, which exceeds the model-implied dispersion on both axes.

### S5. Phase 2 sample-size inputs

The statistical analysis plan specifies the observed variance components in Table S2 as the inputs to the Phase 2 sample-size determination.

The model-implied paired-difference SD for content, 0.939, exceeds the $\leq 0.8$ assumed at design stage and the $\leq 0.6$ that the 0.25 margin requires for $\geq 0.85$ power. Reaching that margin on content in Phase 2 would therefore require a larger sample, a lower dispersion, or both; the sample size required at 0.939 was not computed.

The $\sigma$_rater estimates are weakly identified from three raters (S3) and should be re-estimated on a larger roster before they carry a sample-size calculation. The Phase 1 values were obtained with the current QSI anchors and may not carry over to a refined instrument. Rater calibration against documented agreement targets was not part of the Phase 1 design beyond the training workshop and the eight-case familiarisation round; the agreement estimates in S1 make it a candidate intervention for Phase 2.

### S6. Prior-sensitivity assessment

**Table S6.** Prior-sensitivity by posterior mean difference, 95% credible interval, non-inferiority probability at the 0.5 margin, maximum $\hat{R}$ across model parameters, and divergent-transition count.

| **Prior scale** | **Composite** | $\hat{\Delta}$ | **95% CrI** | **P(Δ > −0.5)** | **Max $\hat{R}$** | **Divergences** |
|---|---|---|---|---|---|---|
| 1.0 (primary) | Content | +0.048 | −0.181 to +0.277 | >0.999 | 1.000 | 0 |
| 1.0 (primary) | Presentation | +0.281 | +0.099 to +0.467 | >0.999 | 1.000 | 0 |
| 2.5 | Content | +0.039 | −0.188 to +0.262 | >0.999 | 1.000 | 0 |
| 2.5 | Presentation | +0.276 | +0.083 to +0.473 | >0.999 | 1.000 | 0 |
| 5.0 (widest) | Content | +0.037 | −0.179 to +0.252 | >0.999 | **1.010** | 0 |
| 5.0 (widest) | Presentation | +0.274 | +0.076 to +0.468 | >0.999 | 1.000 | **6** |

Across a fivefold widening of the prior scales the content estimate moves by 0.011 and the presentation estimate by 0.007, in both cases well inside the credible intervals, and the non-inferiority probability is unchanged at every scale.

The convergence criterion for the primary fits was $\hat{R} < 1.01$ with zero divergent transitions. At the widest prior scale the content refit reached maximum $\hat{R} = 1.010$ with zero divergent transitions and the presentation refit produced six divergent transitions with maximum $\hat{R} = 1.000$; both are reported as diagnostics. No verdict reported in the manuscript depends on either refit.

### S7. Counterbalancing and per-rater checks

Rater codes R1 to R3 do not correspond to author order and are not linkable to individuals outside the controlled study record.

**Table S7a.** Round fixed effect (b_round) from the primary Bayesian model; posterior mean and 95% credible interval.

| Composite | Round effect |
|---|---|
| Content | +0.007 (−0.204 to +0.216) |
| Presentation | −0.096 (−0.287 to +0.092) |
| Overall | −0.050 (−0.208 to +0.109) |

The round effect is consistent with zero on every composite.

**Table S7b.** Arm means and differences by rater. R1 scored 37 cases, R2 19 cases, R3 18 cases.

| Rater | Composite | Manual | AI-assisted | Δ |
|---|---|---|---|---|
| R1 | Content | 4.43 | 4.63 | +0.20 |
| R1 | Presentation | 4.41 | 4.62 | +0.22 |
| R1 | Overall | 4.42 | 4.63 | +0.21 |
| R2 | Content | 3.28 | 2.98 | −0.30 |
| R2 | Presentation | 3.89 | 4.26 | +0.37 |
| R2 | Overall | 3.59 | 3.62 | +0.04 |
| R3 | Content | 3.76 | 3.81 | +0.06 |
| R3 | Presentation | 3.78 | 4.06 | +0.28 |
| R3 | Overall | 3.77 | 3.94 | +0.17 |

The pre-specified check that the contrast is not driven by a single rater is met: every rater scored the AI-assisted arm at or above the manual arm on the overall composite. R2's content and presentation differences are of opposite sign and largely offset, which is why R2's overall difference (+0.04) is the smallest of the three. Absolute score levels differ between raters in both arms, with R1 scoring approximately one point above R2 and R3 on the content composite. The arm contrast is preserved across that offset. The rater term of the hierarchical model absorbs the level difference, and S2 shows it to be the dominant variance component.

The pre-specified outlier rule excludes paired differences exceeding three standard deviations in absolute value. Three pairs met the rule, two on the content composite and one on the presentation composite, and all three originated with R2. The refits are reported in Table 2 of the main text and did not alter the non-inferiority conclusions on either composite. Case identifiers for the excluded pairs are held in the controlled study record.

The 148 eligible scoring rows carried no item-level missingness, so the missing-data refit specified in the analysis plan was not triggered.

### S8. Per-dimension non-inferiority

The dimension-level analysis is a pre-specified secondary and is presented graphically in Figure 3 of the main text. Table S8 gives the Bayesian and frequentist estimates for each dimension.

**Table S8.** Per-dimension non-inferiority at the 0.5 margin. The Bayesian basis is primary; the frequentist linear mixed model on paired differences is reported alongside, with Holm adjustment across the six dimensions [19]. Non-inferiority is concluded on the Bayesian basis where $P(\Delta > -0.5) \geq 0.975$.

| Dimension | Bayesian $\hat{\Delta}$ (95% CrI) | $P(\Delta > -0.5)$ | Bayesian verdict | Frequentist $\hat{\Delta}$ (95% CI) | Raw p | Holm p | Frequentist verdict |
|---|---|---|---|---|---|---|---|
| Accuracy | −0.12 (−0.52 to +0.28) | 0.967 | Not concluded | −0.17 (−0.49 to +0.14) | 0.0205 | 0.0205 | Non-inferior |
| Completeness | +0.07 (−0.26 to +0.43) | 0.999 | Non-inferior | +0.04 (−0.25 to +0.33) | 0.0002 | 0.0005 | Non-inferior |
| Relevance | +0.26 (−0.02 to +0.53) | >0.999 | Non-inferior | +0.24 (−0.02 to +0.50) | <0.0001 | <0.0001 | Non-inferior |
| Organisation | +0.28 (−0.02 to +0.58) | >0.999 | Non-inferior | +0.26 (+0.02 to +0.51) | <0.0001 | <0.0001 | Non-inferior |
| Language | −0.11 (−0.41 to +0.19) | 0.996 | Non-inferior | −0.14 (−0.43 to +0.15) | 0.0073 | 0.0145 | Non-inferior |
| Succinctness | +0.70 (+0.40 to +1.02) | >0.999 | Non-inferior | +0.69 (+0.31 to +1.07) | <0.0001 | <0.0001 | Non-inferior |

The two models disagree only on accuracy, which showed the lowest inter-rater agreement of the nine QSI elements (S1) and the largest score dispersion of the six dimensions; since the frequentist model treats rater as a fixed effect and omits the case-by-arm term (S3), it is the dimension on which a disagreement was expected to appear first.

### Abbreviations

CrI, credible interval; CI, confidence interval; ICC, intraclass correlation coefficient; LMM, linear mixed model; NUTS, No-U-Turn sampler; QSI, Quality Summary Index; $\hat{R}$, Gelman–Rubin convergence diagnostic; sd_d, paired-difference standard deviation.

### References

Reference numbers follow the main manuscript.

10. Creeden J, Olivecrona M, Soriano A. A blinded, counterbalanced rater design for evaluating AI-assisted summarisation of tertiary clinical genomics reports: methodology of the QNOMX-VHIR-CPSP-001 Phase 1 study. medRxiv. 2026. doi:10.64898/2026.06.11.26355467.

17. Hayes AF, Krippendorff K. Answering the call for a standard reliability measure for coding data. Commun Methods Meas. 2007;1(1):77–89. doi:10.1080/19312450709336664

19. Holm S. A simple sequentially rejective multiple test procedure. Scand J Stat. 1979;6(2):65–70.
